# Improving cross-ancestry generalizability of genetic risk prediction for short stature using a meta-polygenic risk score

**DOI:** 10.64898/2025.12.16.25342408

**Authors:** Shihui Peng, Tianyuan Lu

## Abstract

**Background:** Short stature (SS) is associated with adverse clinical, psychosocial, and economic outcomes, and early identification of at-risk individuals may enable timely evaluation and intervention. Polygenic risk scores (PRS) for height offer a promising strategy for SS risk stratification. However, substantial ancestry-related differences in PRS distributions and predictive performance limit equitable clinical translation. Improving cross-ancestry generalizability is therefore essential for reliable and fair implementation.

**Methods:** Using whole-genome sequencing and phenotype data from 371,025 participants in the NIH All of Us Research Program, we calculated five ancestry-specific PRS based on the largest height GWAS to date. Participants were randomly divided into training (20%) and testing (80%) datasets. To mitigate ancestry-related distribution shifts, we residualized height and ancestry-specific PRS on genetic principal components and applied model selection to integrate the residualized ancestry-specific PRS into a meta-polygenic risk score (meta-PRS). Predictive accuracy was evaluated both in the full testing dataset and separately within each ancestry group. We also examined whether a single PRS-based risk threshold is generalizable across diverse ancestries. Sensitivity analyses excluded individuals with known causes of SS.

**Results:** The meta-PRS explained the largest proportion of height variance in the testing dataset and outperformed existing cross-ancestry and ancestry-specific PRS across all ancestry groups, except among individuals of European ancestry, where the European-specific PRS showed marginally higher performance. Each 1-standard-deviation decrease in the meta-PRS was associated with a 3.10-fold increase in the odds of SS (area under the receiver operating characteristic curve = 0.853). The meta-PRS substantially reduced ancestry-related PRS distributional differences and produced a consistent monotonic decrease in SS prevalence across PRS deciles, enabling more stable performance across populations and supporting the use of generalizable risk thresholds. Predictive accuracy was similar when restricting to individuals without known causes of SS, consistent with the largely polygenic nature of unexplained SS.

**Conclusions:** A meta-PRS combining ancestry-specific PRS improves both predictive accuracy and cross-ancestry generalizability of PRS-based SS prediction. By mitigating ancestry-related distributional differences, this framework may support the implementation of generalizable PRS thresholds in risk screening strategies.

## Introduction

Short stature (SS) has significant public health implications. Individuals with SS may face psychosocial challenges, reduced quality of life, and an increased likelihood of health conditions that require medical attention^1,2,3^. Although several treatments exist, such as growth hormone therapy^4^, the financial burden can be substantial^5^. Early identification of individuals at elevated risk for SS is therefore valuable, as timely clinical evaluation and intervention can improve outcomes and reduce long-term disease burden.

Multiple tools have been developed to predict adult height and identify children at risk for SS. Classical methods incorporate current height, growth velocity, parental height, and bone age measured from hand-wrist radiographs, such as the Bayley-Pinneau^6^, Tanner-Whitehouse^7^, and Roche-Wainer-Thissen methods^8^. Although bone-age-based prediction improves accuracy, it requires radiographic imaging^9^, specialized clinical facilities, and expert interpretation, limiting its feasibility for routine or population-level screening. Genetic risk prediction offers a promising complementary approach, given that genotyping or sequencing can now be performed at relatively low cost with high accuracy, and that adult height is among the most heritable complex traits, with up to 80%-90% of phenotypic variation attributable to inherited genetic factors^10,11,12^.

Large-scale genome-wide association studies (GWAS) have enabled the construction of polygenic risk scores (PRS) that aggregate information across multiple genetic variants to quantify genetic predisposition to complex traits. PRS for height are among the most accurate PRS developed to date, capturing a substantial proportion of the genetic contribution to adult height^13,14^. Importantly, prior work has shown that a height PRS can predict future risk of SS in adulthood with an area under the receiver operating characteristic curve (AUROC) above 0.8 among children from the Avon Longitudinal Study of Parents and Children, demonstrating its potential clinical utility^15^. However, most height GWAS, and consequently most PRS, have been developed and evaluated primarily in individuals of European ancestry. When applied to non-European ancestry populations, PRS performance can be substantially reduced, likely due to differences in allele frequencies, linkage disequilibrium patterns, genetic architecture, and environmental or sociocultural factors^16,17,18^. Thus, improving the generalizability of PRS-based SS risk prediction in multi-ancestry settings is important for making optimal use of the existing genetic data and reducing health and healthcare disparities^19^.

In this study, based on 74,205 participants of diverse genetic ancestries from the NIH All of Us (AoU) Research Program^20^, we developed a meta-PRS for adult height by combining ancestry-specific PRS derived from the largest GWAS to date^14^. Using an independent testing dataset (N = 296,820), we evaluated the performance of this meta-PRS relative to the original ancestry-specific PRS and a previously developed cross-ancestry PRS^14^. We examined whether the meta-PRS allows for universal risk thresholding without introducing disparities, thereby providing equitable predictive utility. This work may provide a framework for enhancing PRS predictive performance and generalizability across diverse ancestries and advancing genetically informed screening for short stature.

## Methods

### Data Source and Participants

We conducted our study using data from the AoU (version 8), a nationwide, ongoing biomedical cohort designed to enroll over one million participants from diverse populations across the United States^21^. Among 414,830 individuals with available short-read whole genome sequencing (WGS) data, demographic information, and height measurements, we included 371,025 participants in this analysis. Participants were excluded if their height was recorded in non-centimeter units, if they had outlier height values (more than 2x interquartile range below the first quartile or above the third quartile), if they were not part of a maximally unrelated set (kinship < 0.1 as provided by AoU), or if they had low-quality genomic data (including sample swaps, contamination, or preparation issues identified by AoU). Participants younger than 18 years at their most recent height measurement were also excluded.

The AoU provided inferred genetic ancestries by comparing participants’ genetic data to reference populations from diverse global ancestries^20^. Each participant was assigned to one of the following broad ancestry groups: European, African or African American, East Asian, South Asian, Admixed American, or Middle Eastern.

### Phenotype Definitions

We used the most recent height measurement (in centimeters) for each participant. SS was defined as height less than two standard deviations (2 SD) below the sex-specific mean in the study cohort^22,23^. Sensitivity analyses were conducted using alternative definitions of SS, including height less than 2.25 SD below the sex-specific mean, or the lowest 2.3% of heights within each sex^22,24^. These definitions follow standard clinical and epidemiological criteria for defining SS and do not differentiate by race, ethnicity, or genetic ancestry.

### Genetic Data and Polygenic Scores

PRS for height were generated from the largest GWAS of height conducted by the Genetic Investigation of Anthropometric Traits (GIANT) consortium^14^. This GWAS included approximately 5.4 million individuals across multiple ancestry groups. Ancestry-specific and cross-ancestry fixed-effects inverse variance weighted meta-analyses were performed, encompassing European, African or African American, East Asian, South Asian, and Hispanic ancestries. Importantly, there is no known sample overlap between the cohorts participating in the GIANT consortium and the AoU study cohort. SBayesC^25^ was applied to the results of each of the five ancestry-specific GWAS and the cross-ancestry GWAS meta-analysis to derive variant weights for PRS.

We calculated six PRS for each AoU participant using the published variant weights in European ancestry PRS, African ancestry PRS, East Asian ancestry PRS, South Asian ancestry PRS, Hispanic ancestry PRS, and a PRS derived based on a cross-ancestry meta-analysis (hereinafter referred to as the cross-ancestry PRS). A lower PRS value indicates an increased genetic risk of SS.

### Meta-PRS Construction

We sought to develop a meta-PRS to harness the predictive power of ancestry-specific PRS while improving cross-ancestry generalizability. Individuals in the study cohort were randomly assigned to a training dataset (20%) and a testing dataset (80%). We used a relatively small training dataset because the prediction models do not require extensive parameter tuning. Meanwhile, a larger testing dataset improves the precision and stability of subsequent model evaluation and cross-ancestry comparisons.

Because allele frequencies and linkage disequilibrium patterns differ across ancestries, PRS often exhibit systematic distributional differences between populations that do not reflect true differences in genetic predisposition. Such distribution shifts^26^ can introduce bias when comparing PRS across ancestry groups and may lead to misclassification of risk in one or more ancestry groups if a single risk threshold is applied. To reduce this ancestry-related variation, we separately regressed height and each of the five ancestry-specific PRS on the first ten genetic principal components (PCs). Using the five residualized ancestry-specific PRS as candidate predictors, we then fitted multivariable linear regression models to predict residualized height. Here, EUR, AFR, EAS, SAS, and HIS denote European ancestry PRS, African ancestry PRS, East Asian ancestry PRS, South Asian ancestry PRS, and Hispanic ancestry PRS respectively, and the full model was specified as:

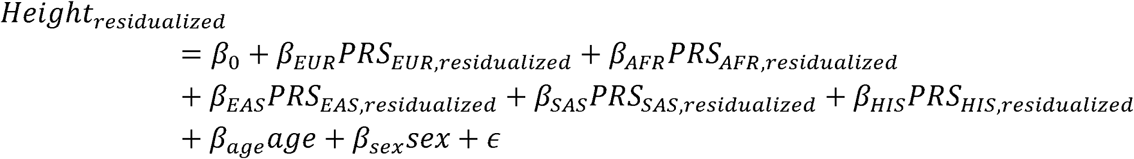

We subsequently fitted nested models by excluding ancestry-specific PRS and calculated the Bayesian Information Criterion (BIC) for each model. We selected the best combination of ancestry-specific PRS, defined as the one minimizing the BIC. This optimized model integrated the selected ancestry-specific PRS into a single combined score (hereinafter referred to as the meta-PRS). Finally, for each individual in the testing dataset, we computed the meta-PRS by applying the coefficients from the selected model to the residualized PRS values obtained using the residualization parameters estimated in the training dataset.

### Performance Evaluation in Testing Dataset

In the testing dataset, we quantified the additional variance in height explained by each PRS (*ΔR*^2^) beyond a baseline model that included age, sex, and the first ten genetic PCs. Specifically, the baseline model was specified as:

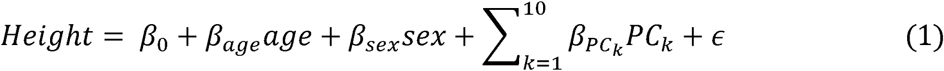

For each PRS (five ancestry-specific PRS, the cross-ancestry PRS, and the meta-PRS), the PRS-inclusive model was specified as:

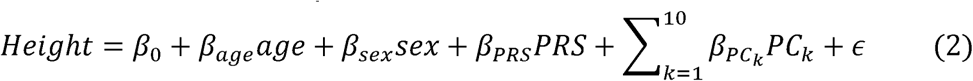

*ΔR*^2^ was computed as

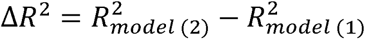

Next, we evaluated the performance of each PRS in identifying individuals with SS. First, we performed logistic regression to estimate the odds ratio (OR) of SS associated with a one-standard deviation decrease in PRS, adjusting for age, sex, and the first ten genetic PCs. We calculated the AUROC and the area under the precision-recall curve (AUPRC), where a non-informative predictor is expected to yield an AUROC of 0.5 and an AUPRC equal to the proportion of cases. These metrics were computed both in the full testing dataset and separately within each genetic ancestry group as well as within females and males. Additionally, we conducted sensitivity analyses using alternative definitions of SS described above.

To examine whether a universal risk threshold could be applied across populations, we compared the distributions of PRS across ancestry groups after min-max normalization. We also assessed the proportions of individuals with SS within each PRS decile. Furthermore, we computed sensitivity, specificity, precision, and balanced accuracy at a series of PRS thresholds (lowest 1%, 2%, 5%, 10%, 15%, 20%, 25%, 30%, 35%, 40%, and 45%), both in the full testing dataset and within each ancestry group. Importantly, these PRS thresholds were defined using the distribution of PRS in the full testing dataset. Thus, each threshold corresponds to a single fixed PRS value that was applied uniformly across all ancestry groups. Individuals with a PRS value below the specified threshold were considered at elevated risk. For these analyses, we focused on comparing the meta-PRS with the European ancestry PRS and the cross-ancestry PRS, as these were the best-performing scores in previous evaluations.

### Sensitivity Analysis

To evaluate whether PRS performance remained robust when individuals with identifiable causes of SS were excluded, we created a subset by removing individuals with Turner syndrome, Noonan syndrome, Prader-Willi syndrome, pituitary dwarfism, achondroplasia, diastrophic dysplasia, growth hormone deficiency, or Down syndrome from the full testing dataset. Prediction metrics, including sensitivity, specificity, precision, and balanced accuracy, were then compared between the original analysis in the full testing dataset and this subset. The meta-PRS and classification thresholds from the full dataset were applied directly.

### Statistical Analysis

All analyses were performed using R (version 4.5.0) within the AoU Researcher Workbench environment.

## Results

### Study Cohort

The study design is illustrated in **Figure 1**. Characteristics of the study cohort are summarized in **Table 1**. Among the 371,025 participants, the mean age (SD) was 53.4 (17.3) years, and 60.2% were females. The average height was 162.7cm in females and 176.5cm in males, and 2% of participants met the definition of SS. The cohort was ancestrally diverse, with the largest group being individuals of European ancestry (57.9%), followed by African or African American ancestry (19.5%), Admixed American ancestry (18.3%), East Asian ancestry (2.5%), South Asian ancestry (1.4%), and Middle Eastern ancestry (0.4%). The training dataset and testing dataset showed nearly identical demographic and anthropometric characteristics (**Table 1** and **Suppl. Table 1**).

**Figure 1.**
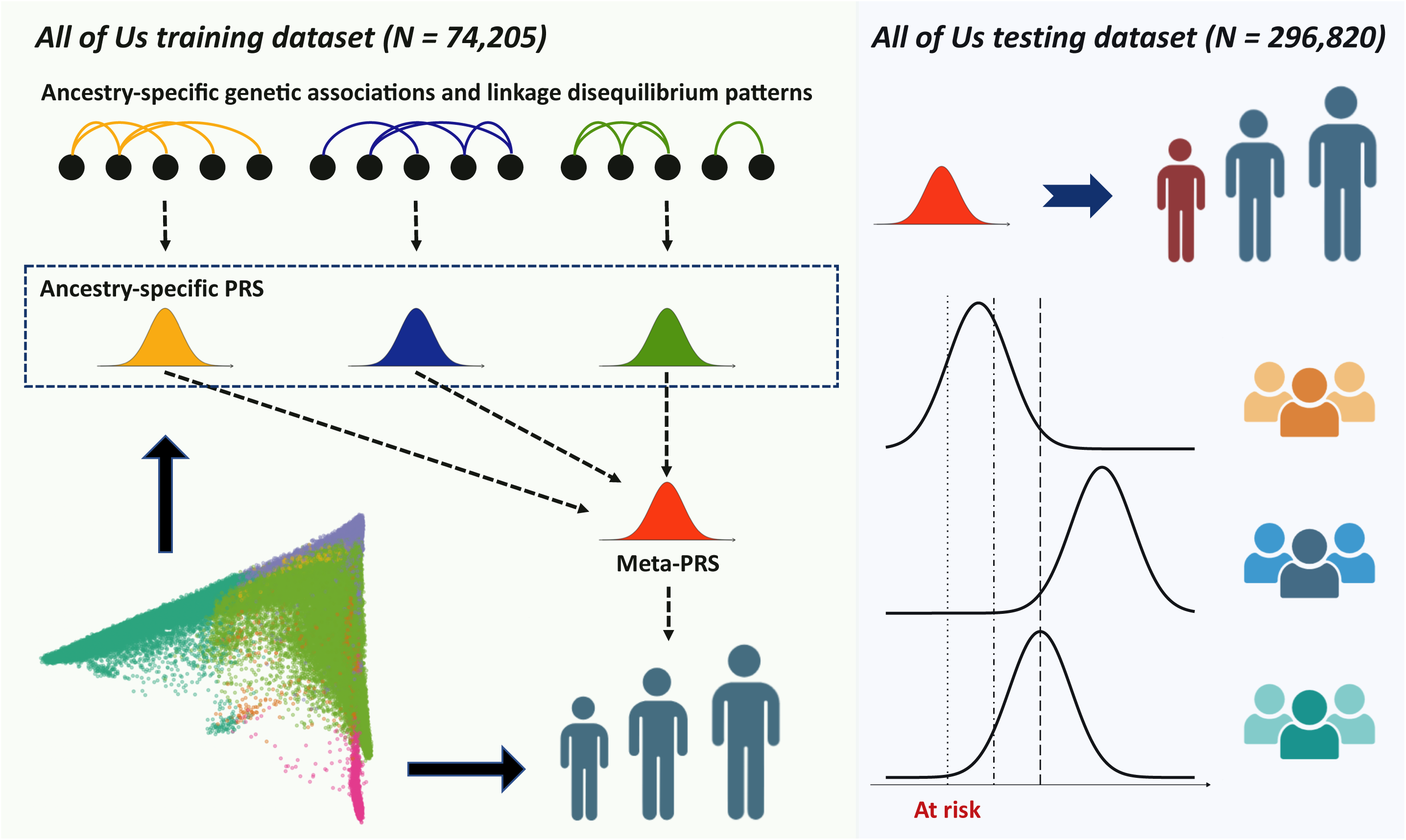
Study design. Participants from the NIH All of Us Research Program were divided into training and testing datasets. Polygenic risk scores (PRS) were calculated using large-scale genome-wide association studies. Population structure acts as a confounder, affecting the distributions of ancestry-specific PRS and height. A meta-PRS was generated by regressing out the effects of the first ten genetic principal components on each ancestry-specific PRS and on height. The performance of PRS was evaluated in the testing dataset, and the generalizability of PRS-based thresholds was assessed across ancestries.

Meta-PRS Construction and Selection of the Optimal PRS

In the training dataset, as expected, each residualized ancestry-specific PRS was significantly associated with residualized height. Due to the correlations among these PRS (**Suppl. Table 2**), model selection based on BIC was performed, resulting in the construction of a meta-PRS as follows:

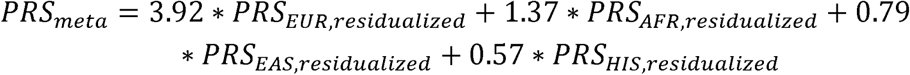

The coefficients for residualizing height and the ancestry-specific PRS are provided in **Suppl. Table 3**. Details of the model selection process are provided in **Suppl. Table 4**.

In the full testing dataset, this meta-PRS achieved a *ΔR*^2^ of 15.11%, outperforming all ancestry-specific PRS as well as the cross-ancestry PRS (9.72%) (**Figure 2**). The European ancestry PRS was the second best, with a *ΔR*^2^ of 14.38%. In ancestry-stratified analyses, the meta-PRS consistently achieved the highest *ΔR*^2^. The only exception was in the European ancestry group, where the European ancestry PRS slightly outperformed the meta-PRS (*ΔR*^2^=18.96% vs 18.76%), though the difference was marginal. Given its strong and consistent performance, we designated the meta-PRS as the optimal and primary score for subsequent analyses.

**Figure 2.**
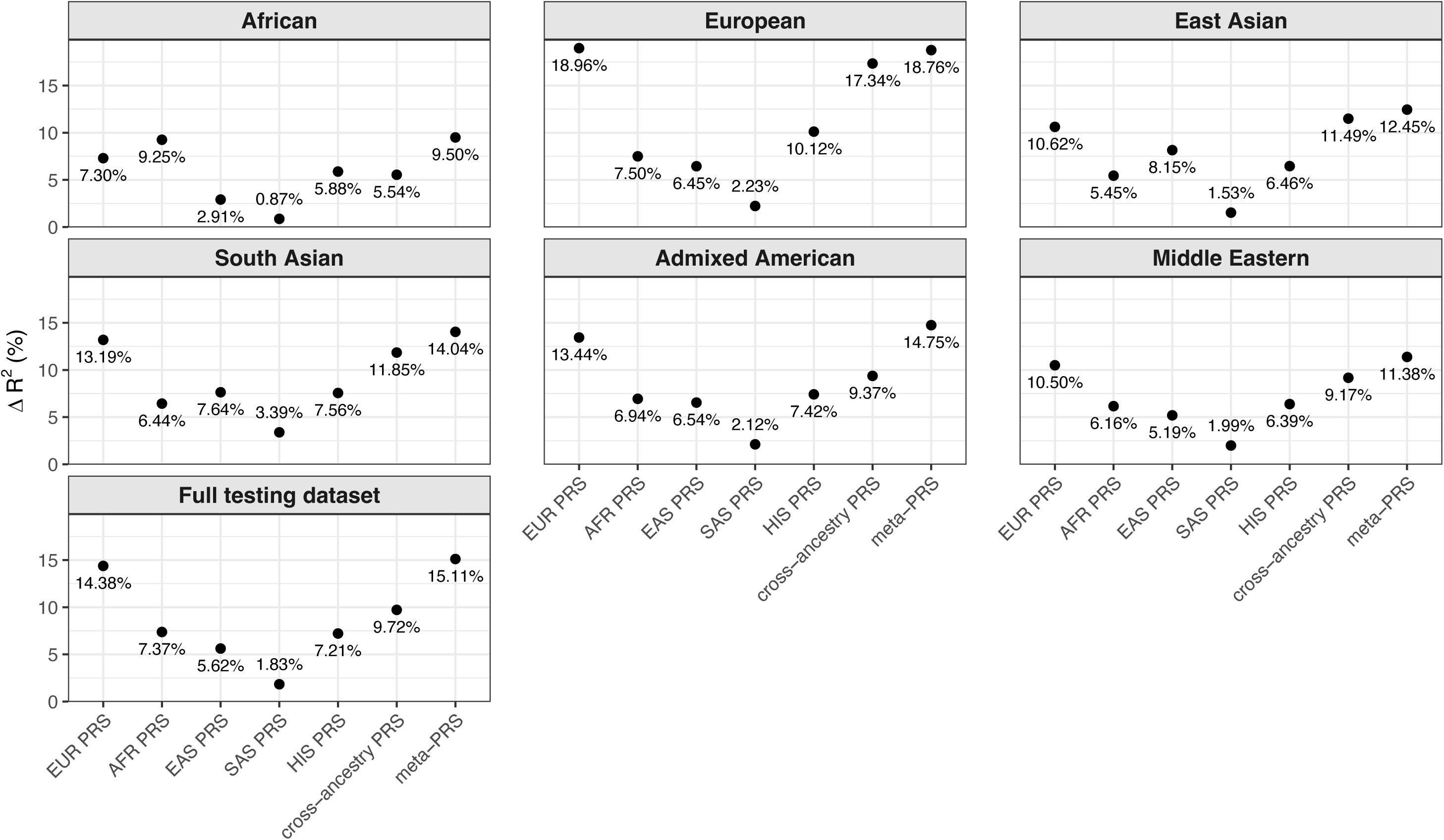
Comparison of predictive performance for height in the testing dataset. Δ*R^2^* quantifies the proportion of variance in height explained by a polygenic risk score (PRS) beyond a baseline model that included age, sex, and the first ten genetic principal components. Δ*R^2^* was calculated for each PRS in both the full testing dataset and within each ancestry group. EUR (European), AFR (African), EAS (East Asian), SAS (South Asian), and HIS (Hispanic) denote the ancestry-specific PRS.

### Meta-PRS Evaluation on SS Prediction

**Figure 3** and **Suppl. Table 5** summarize the predictive performance of the meta-PRS. In the testing dataset, each 1-SD decrease in the meta-PRS for height was associated with a 3.106-fold increase in the odds of SS (95% CI: 3.018–3.196), with an AUROC of 0.853 and an AUPRC of 0.167. Predictive performance varied across ancestries. In the European ancestry group, a 1-SD decrease corresponded to a 4.054-fold increase in SS odds (95% CI: 3.845–4.275), with an AUROC of 0.849 and an AUPRC of 0.101. In contrast, in the African ancestry group, a 1-SD decrease corresponded to a 2.468-fold increase in SS odds (95% CI: 2.311–2.637), with an AUROC of 0.751 and an AUPRC of 0.067. SS prediction did not differ meaningfully between females and males. Sensitivity analyses using alternative definitions of SS yielded consistent results (**Suppl. Table 5**).

**Figure 3.**
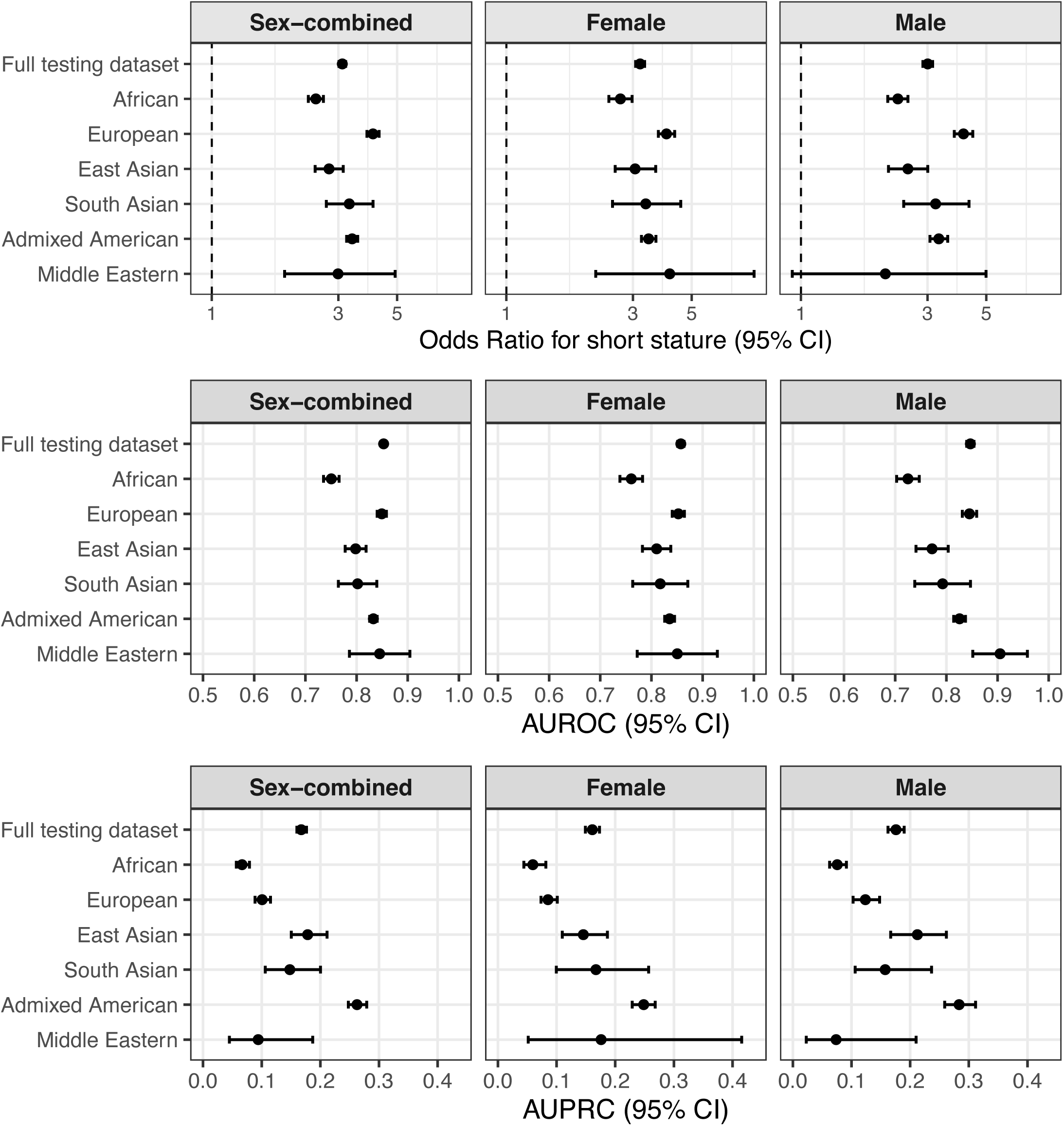
Performance of the meta-polygenic risk score (meta-PRS) in identifying individuals with short stature in the testing dataset. The magnitude of association is expressed as the odds ratio for short stature per one standard deviation decrease in the meta-PRS. Area under the receiver operating characteristic curve (AUROC) and area under the precision-recall curve (AUPRC) were also computed. All metrics were evaluated in both the full testing dataset and within each ancestry group. Sex-stratified analyses were additionally performed. Error bars represent 95% confidence intervals, which for AUROC and AUPRC were estimated using 500 bootstrap samples.

### Distribution of PRS Across Ancestries and Distribution of SS Across PRS Deciles

We observed substantial distribution shifts for both cross-ancestry PRS and EUR PRS across ancestry groups, whereas these shifts were largely mitigated when using the meta-PRS (**Figure 4A**). These distribution shifts also affected the alignment of SS risk across PRS deciles, as the lowest decile of the cross-ancestry PRS and EUR PRS did not consistently capture the highest proportion of SS cases. In contrast, meta-PRS improved the alignment of SS risk. The lowest meta-PRS decile group captured the largest proportion of SS cases in the full testing dataset and within each ancestry-specific subset (**Figure 4B**). Specifically, 8.2% of individuals with a meta-PRS in the lowest decile had SS, compared to only 3.1% of those with a European ancestry PRS and 2.2% of those with a cross-ancestry PRS in the lowest decile. As expected, the proportion of SS decreased monotonically as the meta-PRS increased. In contrast, individuals in the second decile of the cross-ancestry PRS (4.9%) and those in the third decile of the European ancestry PRS (7.2%) were more likely to have SS than those in the respective first deciles.

**Figure 4.**
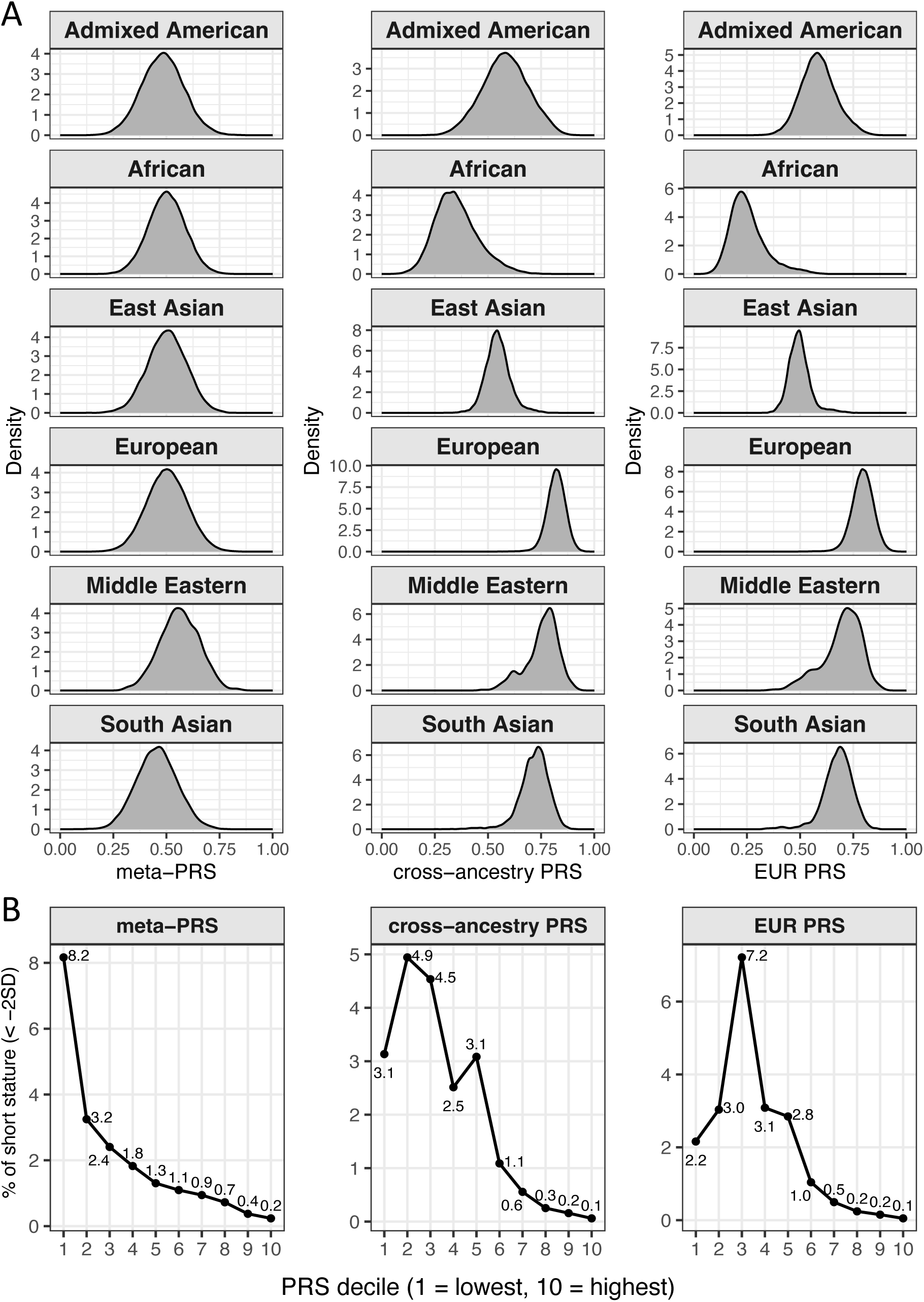
Impacts of PRS distribution shifts in the testing dataset. (A) Density plots of polygenic risk scores (PRS) within each ancestry group. Each PRS was min-max normalized across the full testing dataset for visualization. (B) Distribution of short stature cases across PRS deciles in the full testing dataset. For each PRS, the proportion of individuals with short stature within each decile is shown.

### Performance Metrics Across Thresholds

**Figure 5** and **Suppl. Table 6** display sensitivity, specificity, precision, and balanced accuracy across a wide range of PRS thresholds. The meta-PRS showed stronger cross-ancestry generalizability across all performance metrics compared to the cross-ancestry PRS and European ancestry PRS.

**Figure 5.**
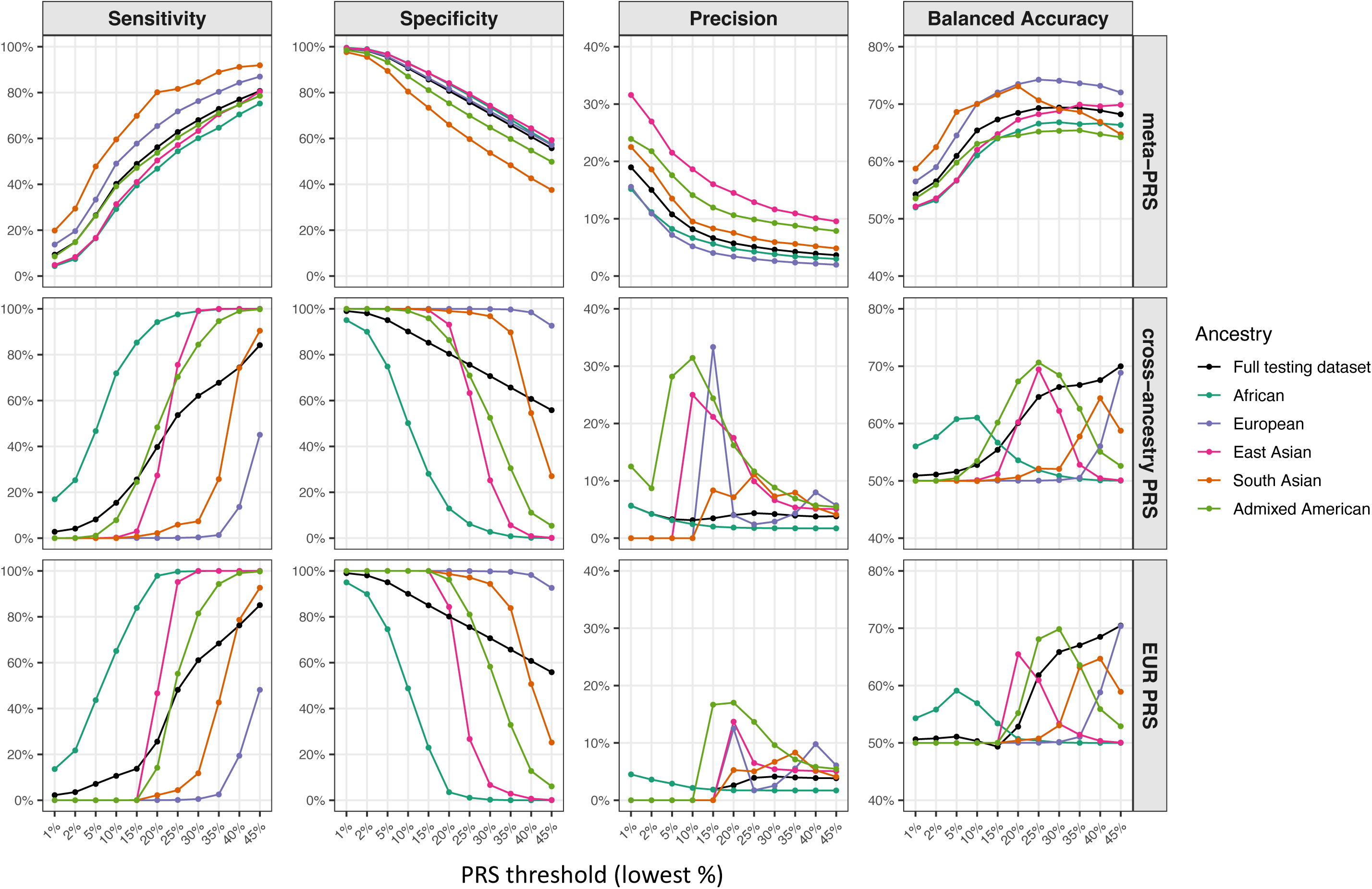
Performance of the meta-polygenic risk score (meta-PRS) for identifying individuals with short stature across specific risk thresholds. The x-axis represents PRS thresholds, with individuals below each threshold classified as predicted short stature cases. Thresholds were determined using the full testing dataset and then applied to each ancestry group. Results for the Middle Eastern ancestry group are not shown due to the limited sample size.

In the full testing dataset, at the 10% threshold, the meta-PRS achieved 40.2% sensitivity, 90.6% specificity, 8.2% precision, and 65.4% balanced accuracy. Across ancestries, sensitivity ranged from 29.2% to 59.6%, specificity from 80.4% to 92.9%, precision from 5.2% to 18.6%, and balanced accuracy from 61.0% to 70.1%. In contrast, although the sensitivity and specificity of cross-ancestry PRS and European ancestry PRS were not substantially lower than those based on the meta-PRS in the full testing dataset, their ancestry-specific performance metrics were highly variable. For the cross-ancestry PRS, sensitivity ranged from 0.0% to 71.9%, specificity from 50.2% to 100.0%, precision from 0.0% to 31.4%, and balanced accuracy from 49.9% to 61.0%. For the European ancestry PRS, sensitivity ranged from 0.0% to 65.1%, specificity from 48.8% to 100.0%, and balanced accuracy from 50.0% to 56.9%. Precision for the European ancestry PRS ranged from 0.0% to 2.2%, consistently lower than that of the meta-PRS.

At the 30% threshold, the meta-PRS achieved 68.0% sensitivity, 70.8% specificity, 4.6% precision, and 69.4% balanced accuracy, with performance metrics remaining more consistent across ancestries. Sensitivity ranged from 60.1% to 84.6%, specificity from 53.7% to 74.3%, precision from 2.6% to 11.6%, and balanced accuracy from 65.3% to 74.0%. Variability in ancestry-specific performance metrics of the cross-ancestry PRS and European ancestry PRS was more pronounced at this threshold. Specifically, for the cross-ancestry PRS, sensitivity ranged from 0.4% to 99.2%, specificity from 2.8% to 99.9%, precision from 1.7% to 8.8%, and balanced accuracy from 50.1% to 68.4%. For the European ancestry PRS, sensitivity ranged from 0.5% to 100.0%, specificity from 0.2% to 99.8%, precision from 1.7% to 9.6%, and balanced accuracy from 50.1% to 69.8%.

### Sensitivity Analysis after Excluding Individuals with Known Causes of SS

After excluding individuals with known causes of SS, 296,687 individuals remained. Applying the meta-PRS to this subset yielded similar performance metrics, with changes in specificity, precision, and balanced accuracy of less than 0.05% across all thresholds in the testing dataset (**Figure 6**). These results were also consistent across ancestries (**Figure 6**).

**Figure 6.**
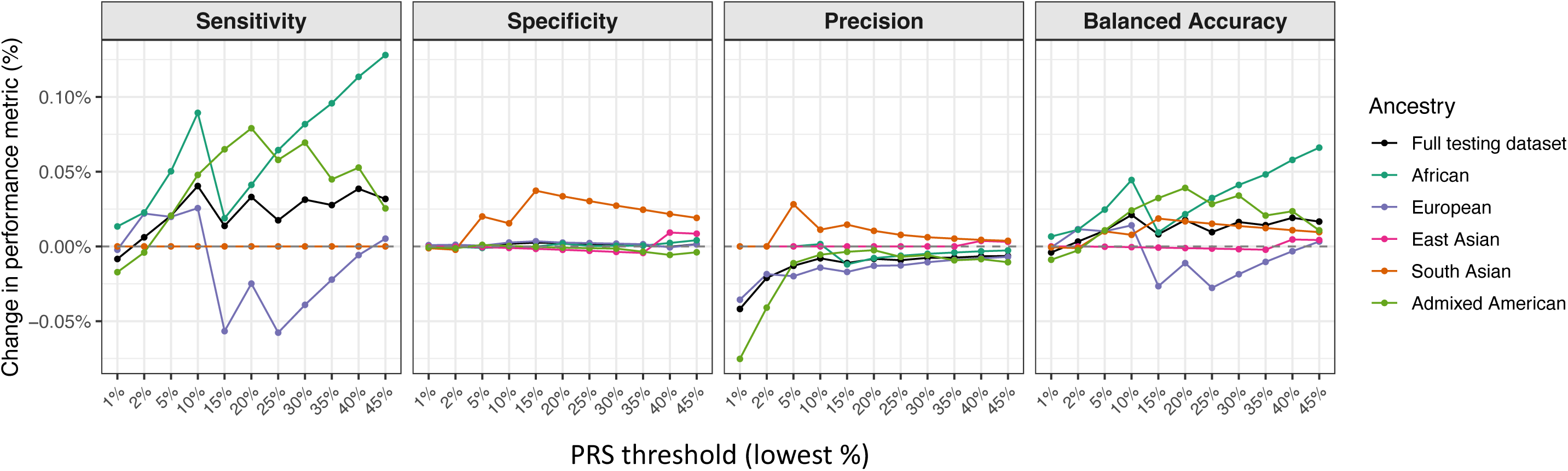
Comparison of performance metrics of the meta-polygenic risk score (meta-PRS) including versus excluding individuals with known causes of short stature. The x-axis represents PRS thresholds, with individuals below each threshold classified as predicted short stature cases. The y-axis shows the difference in prediction metrics, calculated as the values obtained when including all short stature cases minus the values obtained when excluding individuals with known causes of short stature. Results for the Middle Eastern ancestry group are not shown due to the limited sample size.

## Discussion

SS has important clinical, psychosocial, and economic implications, and early identification of at-risk individuals may support timelier evaluation and intervention. In this study, we integrated ancestry-specific PRS derived from the largest height GWAS to date^14^ and used WGS data from participants of diverse ancestries in the AoU to construct a meta-PRS. This meta-PRS increased explained height variance while greatly reducing the ancestry-related distributional differences of PRS. Our findings show that this meta-PRS delivers improved predictive performance across ancestries than existing PRS and may support the use of generalizable thresholds for SS risk prediction and classification.

The meta-PRS strongly predicted both adult height and SS. It explained more variation in adult height than the cross-ancestry PRS and ancestry-specific PRS in the full testing dataset and within each ancestry group, except in the European ancestry group, where it was marginally outperformed by the European ancestry PRS. This performance is expected given the high heritability of height and the availability of multiple well-powered ancestry-specific PRS from large-scale GWAS. Prior studies in other complex traits, such as coronary artery disease^27^, have shown that combining shared genetic effects with ancestry-specific components can improve prediction. Meta-analytic PRS frameworks based on this principle naturally enhance performance by integrating information across ancestries, yielding better prediction than single-ancestry PRS^28,29,30^. We observed minimal differences between predictive accuracy for SS overall and SS without known causes, indicating that the meta-PRS effectively identifies individuals lacking identifiable etiologies. This supports the robustness of the model and aligns with evidence that SS without known causes is largely polygenic^31^. Our findings extend prior work by demonstrating that a meta-PRS framework may help identify individuals at elevated risk for SS without known causes across diverse ancestry groups.

Our results demonstrate that a strong association between the PRS and height or SS in the full testing dataset does not guarantee equitable performance across ancestries. In our study cohort, we observed substantial differences in the distributions of both cross-ancestry PRS and ancestry-specific PRS across ancestry groups. These differences highlight the challenge of establishing a single threshold for clinical use without accounting for population structure. For example, African ancestry participants had systematically lower cross-ancestry PRS and European-derived PRS compared with other groups. Applying a fixed cutoff would classify disproportionately many African ancestry individuals as high genetic risk, reducing specificity, increasing unnecessary clinical follow-up, introducing misclassification, and potentially exacerbating disparities in healthcare access and insurance-related decision-making^17^. Conversely, other ancestry groups would experience reduced sensitivity and precision, leading to under-identification of high-risk individuals and impaired overall predictive performance. These findings underscore the importance of addressing ancestry-specific distributional differences when translating PRS into clinical or screening contexts.

We attempted to address this limitation by intervening directly at the PRS level. Residualizing both height and all ancestry-specific PRS on the first ten genetic PCs before training substantially reduced ancestry-related variation and generated properly weighted inputs for meta-PRS construction. The resulting meta-PRS mitigated distribution shifts and aligned PRS deciles with SS prevalence more consistently than the original scores. This stability is essential for the practical application of equitable thresholds for cross-ancestry risk stratification. Implementing this framework in practice involves two steps. First, new individuals with genetic data should be projected into the same PC space using PC loadings from a reference population, such as All of Us or another cohort^32,33,34^. Second, residualization of the computed ancestry-specific PRS can be performed using the model coefficients obtained from the training dataset, and the residualized PRS can then be used as input to calculate the meta-PRS. While these steps introduce additional computational burden, they are compatible with standard genomic analysis pipelines and enable more equitable and clinically implementable SS risk prediction.

Our study has several limitations. First, we could not apply some of the state-of-the-art methods for creating cross-ancestry PRS, such as PRS-CSx^35^, XPASS^36^, or X-WING^37^, because these methods require full summary statistics, whereas the publicly available GIANT consortium data excluded 23andMe participants^14^. However, based on our results, we believe our proposed framework to improve the generalizability and clinical utility of PRS is widely applicable and expect future studies to revisit our framework using other diverse cohorts once summary statistics of larger GWAS become available. Second, we evaluated major continental ancestry groups, despite substantial heterogeneity within each group, such as the strong diversity among the African populations^38^. Future work may investigate finer-grained subpopulations. Third, our study cohort only included adults. Future work in pediatric or prospective birth cohorts is needed to assess early-life predictive utility. Relatedly, we could not exhaustively account for non-genetic or chronic medical causes of SS during childhood, such as hypothyroidism, chronic kidney disease, or celiac disease, due to the unavailability of data. Finally, in clinical practice, integrating PRS with complementary features, such as parental height, bone-age imaging, endocrine or nutritional markers, or longitudinal growth curves may further improve prediction accuracy. These data were unavailable in the study cohort and warrant incorporation into future multimodal risk prediction models.

In summary, our results show that the meta-PRS explained the greatest proportion of height variance and substantially reduced ancestry-related PRS distributional differences. While differences in predictive performance across ancestry groups remain, this framework represents an important step toward equitable implementation by demonstrating the potential of generalizable thresholds for SS risk prediction across diverse populations. Our findings provide a foundation for developing more inclusive and clinically actionable genetic screening strategies for growth-related outcomes.

## Declarations

### Ethics approval and consent to participate

This research was conducted using secondary data and has been determined to be exempt from IRB review under the University of Wisconsin-Madison Institutional Review Board policies with a formal IRB waiver. De-identified data were obtained from the NIH All of Us Research Program, which implements a broad informed consent model under rigorous ethical and regulatory oversight. The NIH All of Us Research Program collects biospecimens and electronic health records under an IRB-approved protocol that adheres to federal regulations including the Common Rule (45 CFR 46). Participants voluntarily enroll and provide electronic informed consent through a process that includes multimedia materials to enhance understanding, covering topics such as data sharing, privacy protections, and withdrawal rights. Information on this consent process and governance is available at https://allofus.nih.gov/about/protocol/all-us-consent-process.

### Consent for publication

Not applicable.

### Availability of data and materials

Access to the All of Us Research Program is governed by the program’s data access policies and is available to registered researchers in accordance with those guidelines.

### Authors’ contributions

T.L. conceptualized and supervised the study and acquired the data. S.P. and T.L. developed the methodology. S.P. performed the formal analysis, created the visualizations, and drafted the initial manuscript. T.L. and S.P. critically revised the manuscript and approved the final submitted version.

## Data Availability

Access to the All of Us Research Program is governed by the program data access policies and is available to registered researchers in accordance with those guidelines.

## Acknowledgements

We gratefully acknowledge All of Us participants for their contributions, without whom this research would not have been possible. We also thank the National Institutes of Health’s All of Us Research Program for making available the participant data examined in this study. We also acknowledge support from the Interdisciplinary Biological and Health Sciences Consortium at the University of Wisconsin-Madison School of Medicine and Public Health for S.P.’s doctoral training.

