## Supplementary material for "Improving cross-ancestry generalizability of genetic risk prediction for short stature using a meta-polygenic risk score": Table 1

Table 1. Cohort Characteristics. EUR: European ancestry; AFR: African or African American ancestry; EAS: East Asian ancestry; SAS: South Asian ancestry; AMR: Admixed American ancestry; MID: Middle Eastern ancestry.

| **Characteristics** | **All samples** | **Training dataset** | **Testing dataset** | **EUR** | **AFR** | **EAS** | **SAS** | **AMR** | **MID** |
| --- | --- | --- | --- | --- | --- | --- | --- | --- | --- |
| **N (%)** | 371,025 (100.0) | 74,205 (20.0) | 296,820 (80.0) | 214,992 (57.9) | 72,243  (19.5) | 9,286 (2.5) | 5,103 (1.4) | 67,999 (18.3) | 1,402 (0.4) |
| **Age, mean (SD)** | 53.4 (17.3) | 53.4 (17.3) | 53.4 (17.3) | 57.3 (17.2) | 50.9 (15.2) | 45.4 (17.6) | 42.4 (16.4) | 45.8 (15.9) | 46.3 (17.8) |
| **Female, n (%)** | 223,462 (60.2) | 44,623 (60.1) | 178,839 (60.3) | 127,999 (59.5) | 41,886 (58.0) | 5,844 (62.9) | 2,696 (52.8) | 44,294 (65.1) | 743 (53.0) |
| **Height, mean (SD) (cm)** |  |  |  |  |  |  |  |  |  |
| **Female** | 162.7 (7.1) | 162.7 (7.1) | 162.7 (7.1) | 163.8 (6.9) | 163.7 (7.0) | 159.0 (6.4) | 160.4 (6.4) | 159.5 (6.8) | 161.1 (6.6) |
| **Male** | 176.5 (7.7) | 176.5 (7.7) | 176.5 (7.7) | 177.7 (7.3) | 176.8 (7.8) | 171.5 (7.1) | 173.8 (7.3) | 172.8 (7.7) | 175.2 (6.8) |
| **Short stature, n (%)** | 7,467 (2.0) | 1,435 (2.0) | 6,032 (2.0) | 2,107  (1.0) | 1,186  (1.6) | 469 (5.1) | 169 (3.3) | 3,508 (5.2) | 28 (2.0) |
